# Predictive Value of Serum Uric Acid/Serum Creatinine Ratio on Cardiovascular and All-Cause Mortality in Patients with Normal Renal Function: an Analysis from the Real Clinical Practice in Italy

**DOI:** 10.64898/2025.12.11.25342114

**Authors:** Luca Degli Esposti, Melania Leogrande, Valentina Perrone, Margherita Andretta, Marcello Bacca, Antonella Barbieri, Fausto Bartolini, Alessandro Brega, Alessandro Chinellato, Mariarosaria Cillo, Francesco Coria, Stefania Dell’Orco, Fulvio Ferrante, Lotita Gallo, Stefano Grego, Andrea Marinozzi, Cataldo Procacci, Davide Re, Maria Cappuccilli, Claudio Borghi

## Abstract

**BACKGROUND:** Serum uric acid (SUA) has been associated with cardiovascular (CV) and all-cause mortality; however, its interpretation is influenced by renal function. The serum uric acid to creatinine ratio (SUA/sCr) has been proposed as a refined marker, integrating urate burden with renal excretory capacity. Evidence from real-world populations with preserved renal function remains limited.

**METHODS:** A retrospective observational analysis was performed using administrative and laboratory databases from nine Italian healthcare entities, covering more than 7.6 million individuals. Eligible subjects were 18–95 years old, with estimated glomerular filtration rate (eGFR) ≥60 mL/min/1.73 m², serum creatinine (sCr) ≤1.8 mg/dL, and no prior urate-lowering therapy. The index date was defined as the first SUA measurement between January 2017 and March 2022. A total of 739,950 participants were stratified into quintiles of SUA/sCr ratio. Outcomes were two-year all-cause mortality and CV events, including ischemic heart disease, heart failure, cerebrovascular disease, and coronary revascularization.

**RESULTS:** All-cause mortality exhibited a U-shaped distribution, with the lowest risk in mid-quintiles and the highest in extreme quintiles (6.2% in quintile 1, 7.0% in quintile 5). Cardiovascular events showed a progressive increase, from 9.9% in quintile 1 to 11.6% in quintile 5. Sex-stratified analyses indicated higher absolute risks in men, whereas women demonstrated steeper relative increases across higher quintiles. In individuals with hyperuricemia, mortality and CV events rose consistently with increasing SUA/sCr values.

**CONCLUSIONS:** The SUA/sCr ratio is an independent predictor of short-term mortality and cardiovascular events in individuals with preserved renal function. Its incorporation into routine risk stratification may support earlier identification of high-risk patients, particularly those with elevated SUA.

**CLINICAL PERSPECTIVE:** *What Is New?:* In this large, real-world study of nearly 740,000 adults with preserved renal function, the serum uric acid to serum creatinine (SUA/sCr) ratio emerged as a strong, independent predictor of two-year all-cause and cardiovascular mortality. The prognostic value of SUA/sCr was particularly evident in patients with hyperuricemia, where higher quintiles were associated with progressively increased risk of death and cardiovascular events.

*What Are the Clinical Implications?:* The SUA/sCr ratio, based on two inexpensive and widely available laboratory parameters, provides a practical tool for early cardiovascular risk stratification in patients without overt kidney disease.

## INTRODUCTION

Uric acid is the final oxidation product of purine metabolism in humans, and its systemic concentration is determined by the delicate balance between purine intake, endogenous production, renal and gastrointestinal excretion.^1-5^ Although the pathophysiological role of chronic hyperuricemia was initially confined to gout and renal disorders, a growing body of evidence over the past decades has expanded its relevance, positioning serum uric acid (SUA) as a potential independent risk factor for the development of hypertension, metabolic syndrome, chronic kidney disease (CKD), and cardiovascular (CV) disorders.^6-9^

As early as the 1970s, epidemiological studies began to explore the association between SUA and CV outcomes, laying the foundation for ongoing research into its clinical utility in both general and high-risk populations.^10^

In recent years, the Uric Acid Right for Heart Health (URRAH) project, coordinated by the Italian Society of Hypertension, was established to clarify the prognostic significance of SUA and provided robust, large-scale, population-based insights to this field. In particular, the URRAH investigators have consistently demonstrated the role of SUA as an independent predictor of CV and all-cause mortality, even in individuals without overt CV disease.^11^ The initial analyses aimed to determine SUA thresholds that predict CV events and mortality with real-world data from over 23,000 individuals.^12^ Successive URRAH studies identified precise SUA cut-off values associated with fatal myocardial infarction (≥5.70 mg/dL in men, ≥5.26 mg/dL in women),^13^ and with all-cause and CV mortality over a 20-year span.^14^ These results were confirmed by another study involving a Chinese population and carried out in subjects with increasing levels of serum urate without any additional cardiovascular risk factor.^15^

One recent publication from the URRAH project revealed that SUA values ≥5.6 mg/dL were associated with an increased risk of all-cause and CV mortality even in patients without overt comorbidities, emphasizing the need for early identification of at-risk individuals.^16^

Importantly, novel ways to contextualize SUA were also explored by indexing it to renal function. Since SUA levels are substantially influenced by glomerular filtration, the exclusive consideration of SUA without accounting for renal function may reduce its prognostic interpretation. Serum creatinine (sCr), a well-established marker of renal function, is widely available and cost-effective, and has been proposed as a denominator in the serum uric acid to serum creatinine (SUA/sCr) ratio to enhance the discriminatory power of SUA-related metrics.^17,18^ In particular, elevated values of the SUA/sCr ratio identify subjects with overproduction of serum urate within the hyperuricemic population, with a significant increase in the probability of detecting individuals with overactivation of circulating/tissue xanthine oxidase. Previous studies have shown SUA/sCr to be significantly associated with a dysregulation of metabolic, including those with metabolic syndrome^19,20^ and insulin resistance.^21^ Furthermore, elevated values of SUA/sCr ratio have been linked to worse CV outcomes across several clinically relevant subgroups, such as individuals with type 2 diabetes,^22^ postmenopausal women,^20,23^ and patients affected by CKD,^24,25^ suggesting that this index may reflect an interplay between metabolic and renal determinants of CV risk.

The URRAH project evaluated a cohort of 20,724 Italian participants followed for up to 240 months, and identified a prognostic cut-off value of SUA/sCr >5.35, which was consistent across men and women. Individuals with a ratio above this threshold had a significantly higher risk of CV events, with hazard ratios of 1.159 in the total population, 1.161 in men, and 1.444 in women. The risk increased further in higher SUA/sCr quintiles.^17^ The same evidence was conformed in maintenance hemodialysis patients, where higher levels of SUA/SCr are associated with a greater risk of cardiovascular mortality thereby supporting the primary role of urate overproduction.

The present analysis aims to evaluate the prognostic relevance of SUA/sCr ratio in predicting two-year all-cause mortality and CV events in individuals with preserved renal function, with a particular focus on sex-based differences in risk patterns. For this purpose, the variations in mortality and CV event rates with their distinct trajectories in men and women were investigated across increasing quintiles of the SUA/sCr ratio.

## METHODS

### Study Design and Data Source

An observational retrospective analysis was conducted by integrating administrative and laboratory databases of Italian healthcare entities geographically distributed across the national territory, including a population of over 7.6 million health-assisted individuals. Administrative databases are extensive data repositories containing information on medical services and resources reimbursed by the Italian National Health System (INHS), which provides universal healthcare coverage to all citizens and legal foreign residents since 1978. Hence, all individuals residing in the catchment area of healthcare entities included in this analysis can be tracked within these administrative flows. Specifically, the following databases were browsed: i) beneficiaries’ database, to get information on patients’ demographic information, including sex, age, and date of death; ii) pharmaceuticals database, containing details on medications reimbursed by the INHS, such as Anatomical Therapeutic Chemical (ATC) codes, number of packages, units per package, unit cost, and prescription dates; iii) hospitalization database, to capture data on hospital admissions, including discharge diagnosis codes classified according to the International Classification of Diseases, Ninth Revision, Clinical Modification (ICD-9-CM), Diagnosis Related Groups (DRGs), and corresponding DRG-based charges covered by the INHS; iv) outpatient specialist services database, documenting information on specialist visits and diagnostic tests, including delivery dates, activity descriptions, and related charges for laboratory tests or specialist visits; v) exemption database, which records active payment waiver codes that allow patients to receive services or treatments free of charge due to specific medical conditions; vi) laboratory database, to collect SUA measurement, and information on renal function indexes, namely sCr and estimated glomerular filtration rate (eGFR).^26^ The dataset used consists solely of anonymized data. All the results of the analyses were produced and presented as aggregated summaries. Approval has been obtained from the ethics committees of the participating healthcare entities.

### Study Population

From January 2017 to March 2022, subjects with available SUA, sCr and eGFR values were screened and then those meeting the following criteria were included: age between 18 and 95 years; eGFR ≥60 mL/min/1.73 m²; sCr ≤1.8 mg/dL; no current or prior treatment with urate-lowering therapy (ATC code M04AA); no history of dialysis treatment.

The index-date corresponded to the time of the first available SUA determination. A 1-year period prior to index-date was used to describe patients’ previous history (characterization period). The follow-up period (at least 2 years) began at index-date and ended at the end of study period, or patient’s death, or exiting the database (whichever occurred first).

### Patients’ Demographic and Clinical Characteristics

Age was recorded at index-date and reported as mean ± standard deviation (SD), sex was presented as proportion of males.

Clinical comorbidities were evaluated using the Charlson Comorbidity Index (CCI), a validated scoring system incorporating 19 weighted conditions associated with mortality risk.^27^ Comorbid conditions were identified through pharmacological treatments and hospital admissions recorded within the 12 months preceding the index-date.

### Association of SUA/Scr Ratio with Clinical Outcomes

The analysis evaluated the potential association of SUA/sCr ratio with the following outcomes: (A) all-cause mortality; (B) cardiovascular and cerebrovascular events, evaluated as ischemic heart diseases, heart failure, cerebrovascular conditions, or by the presence of procedures of cardiac revascularization and other invasive coronary interventions (Table S1 of the Supplementary materials).

### Statistical Analysis

Continuous variables are presented as mean ± SD, and median values, while categorical variables are expressed as numbers and percentages. The variable SUA/sCr ratio was divided into five increasing quintiles of 147,990 subjects each, to be used in purely descriptive statistics. A focused analysis was conducted in a subset of patients with SUA levels above the upper reference value (7 mg/dL for males and 6 mg/dL for females). All the statistical analyses were performed using STATA SE version 17.0 (StataCorp LLC, College Station,TX, USA).

## RESULTS

From a sample of 7,629,514 Italian citizens, 739,950 subjects met the inclusion criteria. After calculating the SUA/sCr ratios for all participants, individuals were ranked in ascending order based on their values. The entire population was then divided into five equal-sized groups, or quintiles, each including 147,990 individuals. These quintiles represent progressively increasing levels of SUA/sCr ratios, with the first quintile including the lowest 20% and the fifth quintile representing the highest 20%. As Table 1 shows, the SUA/sCr ratio was by 4.3 (±0.6) in the first quintile, progressively rising to 5.4 (±0.2), 6.2 (±0.2), 7.1 (±0.3), and reaching 9.1 (±2.5) in the highest quintiles. Age remained relatively stable across the quintiles, with a slight downward trend: 59.0 years (±17.7) in the first quintile, followed by 58.3 (±17.8), 58.4 (±17.7), 58.5 (±17.8), and 58.8 (±18.0) years in the fifth.

**Table 1.** Demographic, biochemical and clinical characteristics of the overall patients divided by SUA/sCr quintiles.

| Quintiles<br>SUA/sCr | Number of<br>patients | Uric<br>acid(mg/dL),<br>mean ± SD | eGFR (mL/min/1.73<br>m<sup>2</sup>),<br>mean ± SD | sCr (mg/dL),<br>mean ± SD | SUA/sCr,<br>mean ± SD | Age (years),<br>mean ± SD | Male sex,<br>N (%) | CCI,<br>mean ± SD |
| --- | --- | --- | --- | --- | --- | --- | --- | --- |
| 1 | 147,990 | 3.7 ± 0.9 | 88.2 ± 20.4 | 0.9 ± 0.2 | 4.3 ± 0.6 | 59.0 ± 17.7 | 69,612 (47.0%) | 0.4 ± 0.8 |
| 2 | 147,990 | 4.6 ± 0.9 | 91.4 ± 20.4 | 0.8 ± 0.2 | 5.4 ± 0.2 | 58.3 ± 17.8 | 74,546 (50.4%) | 0.3 ± 0.7 |
| 3 | 147,990 | 5.0 ± 1.0 | 95.5 ± 22.0 | 0.8 ± 0.2 | 6.2 ± 0.2 | 58.4 ± 17.7 | 72,589 (49.0%) | 0.3 ± 0.7 |
| 4 | 147,990 | 5.4 ± 1.1 | 100.5 ± 23.8 | 0.8 ± 0.2 | 7.1 ± 0.3 | 58.5 ± 17.8 | 69,358 (46.9%) | 0.4 ± 0.8 |
| 5 | 147,990 | 6.0 ± 1.3 | 115.9 ± 61.9 | 0.7 ± 0.2 | 9.1 ± 2.5 | 58.8 ± 18.0 | 56,695 (38.3%) | 0.4 ± 0.8 |

Sex distribution had a varying pattern, as the proportion of male patients began at 47.0% (69,612 individuals) in the first quintile, rose slightly to 50.4% in the second, then dropped to 49.0% in the third, 46.9% in the fourth, and then decreased more sharply to 38.3% (56,695) in the fifth quintile. The CCI was quite stable, ranging between 0.3 and 0.4 across all quintiles, with slight fluctuations (0.4 in the first and fifth quintiles, 0.3–0.4 in the others).

In the overall population two-year all-cause mortality followed a u-shaped pattern, as it started at 6.2% (9,163 deaths) in the first quintile declined to 4.3% in the second and 4.2% in the third quintile, respectively, then slightly increased to 4.6% in the fourth, and reached a zenith at 7.0% (10,309 deaths) in the fifth (Figure 1A). The percentage of patients experiencing CV events within two years displayed a similar increasing trend, starting at 9.9% (14,712 patients) in the first quintile, followed by a slight decline to 8.7% in the second, then climbing to 9.0%, 9.5% in the third and fourth, and reaching 11.6% (17,233 patients) in the fifth quintile (Figure 1B).

**Figure 1.**
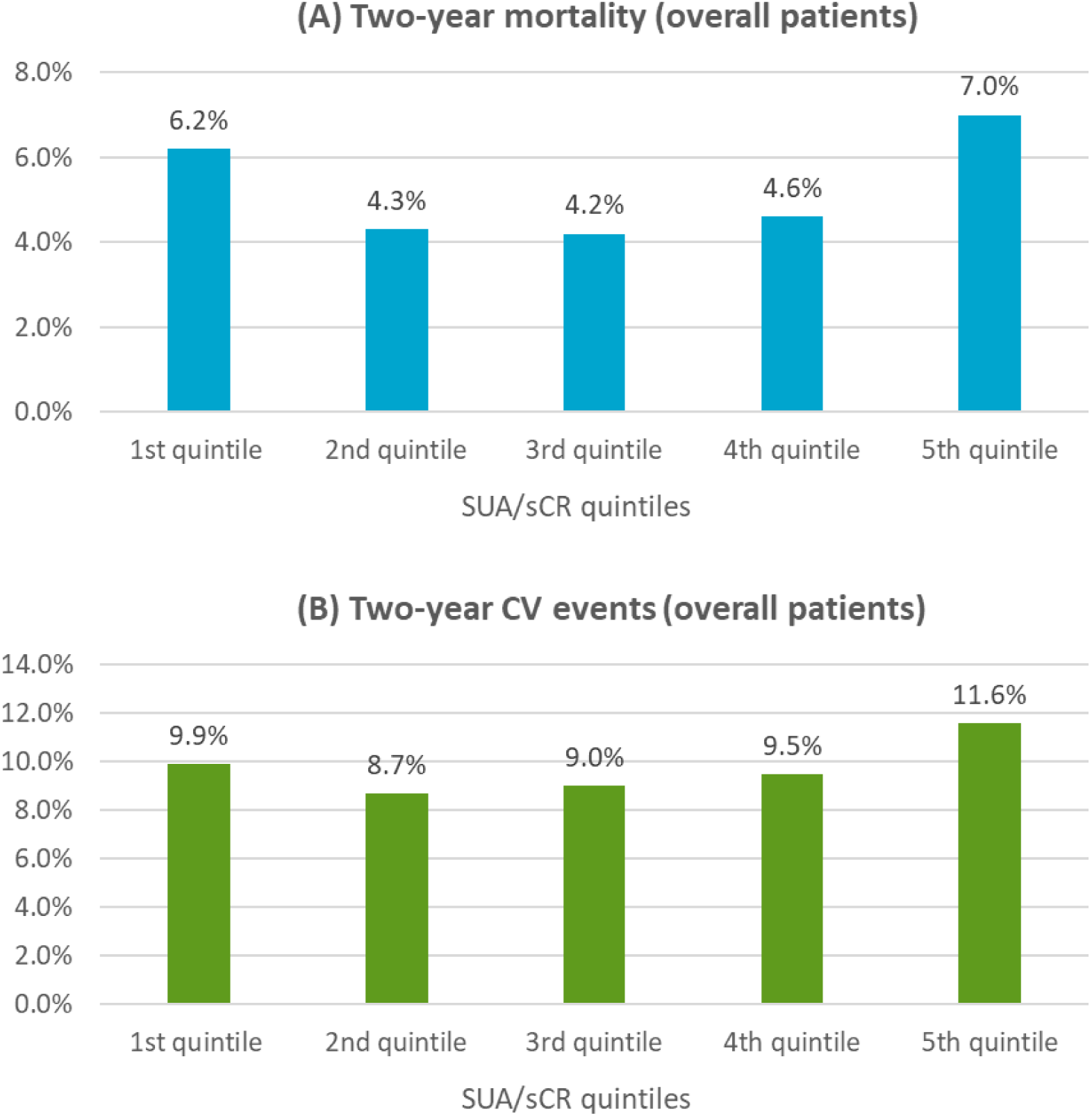
Two-year mortality **(A)** and CV events **(B)** in the overall patients divided by SUA/sCr quintiles.

When stratifying patients by sexes (Table 2), age showed a decreasing trend in men, from 61.5 years in the lowest quintile to 57.6 years in the highest. Among women, age increased slightly from 56.8 to 59.6 years across the same range. Hence, men in lower quintiles tend to be older than women, but this difference narrows in higher quintiles.

**Table 2.** Demographic, biochemical and clinical characteristics of the overall patients divided by SUA/sCr quintiles and by sex.

| Quintiles<br>SUA/sCr | Number of<br>males | Uric acid (mg/dL),<br>mean ± SD | eGFR (mL/min/1.73<br>m<sup>2</sup>),<br>mean ± SD | sCr (mg/dL),<br>mean ± SD | SUA/sCr,<br>mean ± SD | Age (years),<br>mean ± SD | CCI,<br>mean ± SD |
| --- | --- | --- | --- | --- | --- | --- | --- |
| 1 | 69,612 | 4.2 ± 0.9 | 86.9 ± 19.7 | 1.0 ± 0.2 | 4.3 ± 0.6 | 61.5 ± 17.2 | 0.4 ± 0.9 |
| 2 | 74,546 | 5.1 ± 0.8 | 90.7 ± 18.5 | 0.9 ± 0.1 | 5.4 ± 0.2 | 59.1 ± 17.3 | 0.4 ± 0.8 |
| 3 | 72,589 | 5.6 ± 0.9 | 95.3 ± 19.7 | 0.9 ± 0.1 | 6.2 ± 0.2 | 58.4 ± 17.2 | 0.4 ± 0.8 |
| 4 | 69,358 | 6.1 ± 0.9 | 101.6 ± 20.7 | 0.9 ± 0.1 | 7.1 ± 0.3 | 57.7 ± 16.9 | 0.4 ± 0.8 |
| 5 | 56,695 | 6.8 ± 1.2 | 117.4 ± 44.3 | 0.8 ± 0.1 | 8.9 ± 1.6 | 57.6 ± 16.4 | 0.4 ± 0.9 |
| Quintiles<br>SUA/sCr | Number of<br>females | Uric acid (mg/dL),<br>mean ± SD | eGFR (mL/min/1.73<br>m<sup>2</sup>),<br>mean ± SD | sCr (mg/dL),<br>mean ± SD | SUA/sCr,<br>mean ± SD | Age (years),<br>mean ± SD | CCI,<br>mean ± SD |
| 1 | 78,378 | 3.2 ± 0.7 | 89.4 ± 20.9 | 0.7 ± 0.1 | 4.3 ± 0.6 | 56.8 ± 17.9 | 0.3 ± 0.7 |
| 2 | 73,444 | 4.0 ± 0.6 | 92.2 ± 22.2 | 0.7 ± 0.1 | 5.4 ± 0.2 | 57.5 ± 18.2 | 0.3 ± 0.7 |
| 3 | 75,401 | 4.4 ± 0.7 | 95.7 ± 23.9 | 0.7 ± 0.1 | 6.2 ± 0.2 | 58.4 ± 18.3 | 0.3 ± 0.7 |
| 4 | 78,632 | 4.8 ± 0.8 | 99.6 ± 26.3 | 0.7 ± 0.1 | 7.1 ± 0.3 | 59.2 ± 18.4 | 0.4 ± 0.8 |
| 5 | 91,295 | 5.6 ± 1.2 | 114.9 ± 70.7 | 0.6 ± 0.1 | 9.2 ± 2.8 | 59.6 ± 18.9 | 0.4 ± 0.8 |

The CCI confirmed to be relatively stable in both groups, fluctuating modestly between 0.3 and 0.4, suggesting comparable comorbidity burdens across quintiles and sexes.

The percentage of deaths within two years from the first SUA determination decreased in middle quintiles before rising again in both sexes. In males, mortality dropped from 8.3% in the first quintile to 4.8–5.0% in the middle groups, then rose up to 7.4% in the highest quintile (Figure 2A). In females, a similar pattern was observed, from 4.3% in the lowest to 3.5–4.3% in the middle, rising to 6.7% in the highest quintile (Figure 2B).

**Figure 2.**
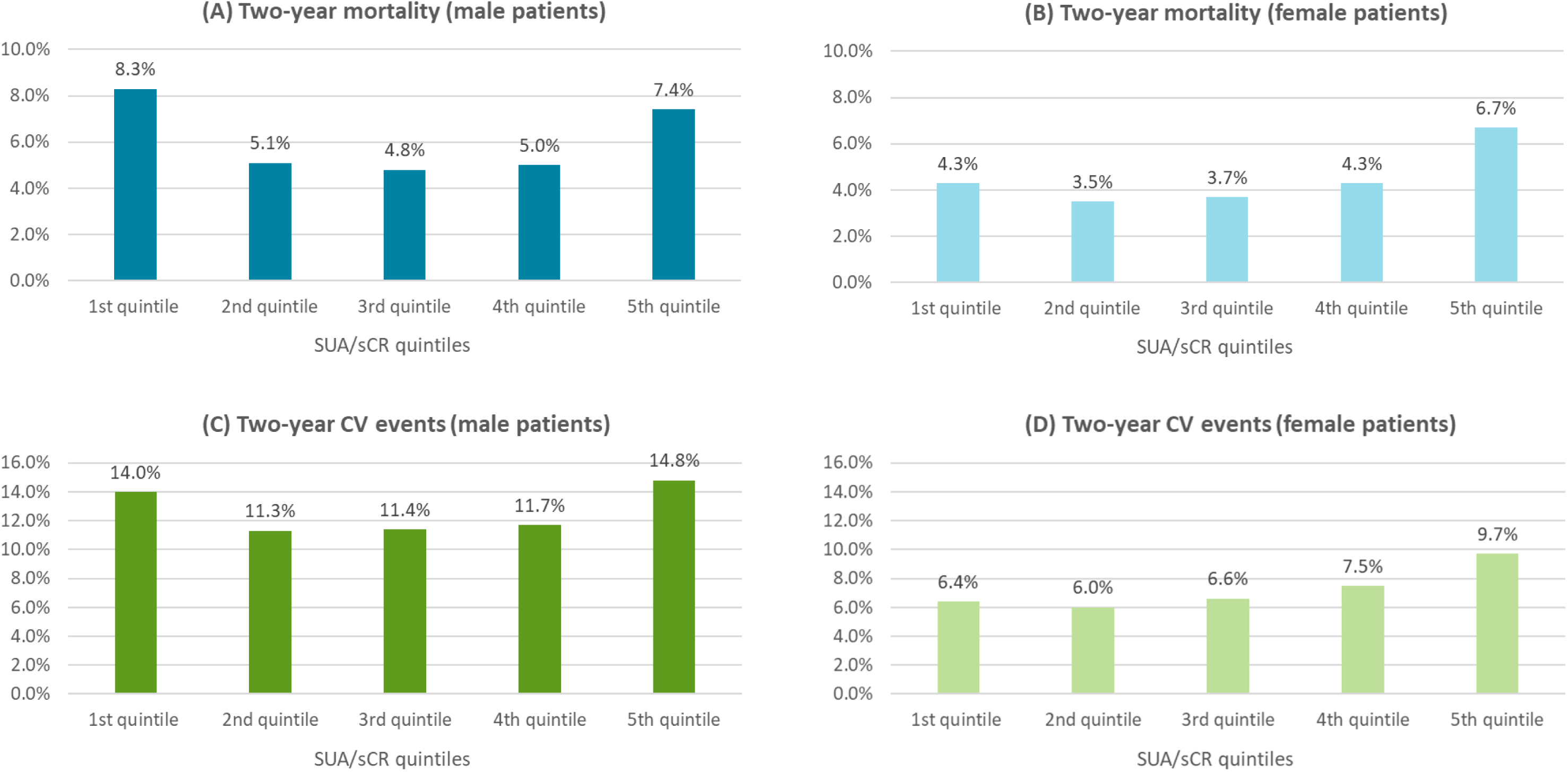
Two-year mortality in males **(A)** and in females **(B)**; CV events in males **(C)** and in females **(D)** divided by SUA/sCr quintiles.

CV events within two years from the first SUA determination followed an upward trend in both sexes. Men showed an increase from 14.0% in the lowest quintile to 14.8% in the highest, with a dip to 11.3–11.7% in the intermediate groups (Figure 2C). Women start much lower, at 6.4%, and steadily climb to 9.7% in the highest quintile (Figure 2D).

To exclude the possibility that the described U-shaped pattern is the consequence of reduced serum urate due to underlying clinical conditions, a secondary analysis was carried out in the subpopulation of patients with absolute serum urate level above the “normal” value according to traditional classification of hyperuricemia.

Table 3 shows the demographic, biochemical, and clinical characteristics of subjects with SUA values above the upper reference threshold (> 7.0 mg/dL in males and > 6.0 mg/dL in females), categorized by quintiles of SUA/sCr ratio. Among males (N=35,425), SUA levels increased moderately from 7.3 ± 0.3 mg/dL in the lowest quintile to 8.2 ± 1.2 mg/dL in the highest, while sCr declined from 1.2 ± 0.1 mg/dL to 0.8 ± 0.1 mg/dL. As a result, the SUA/sCr ratio rose from 6.4 ± 0.4 to 10.6 ± 1.9 across quintiles. A similar trend was observed in females (N=34,335), with SUA increasing from 6.3 ± 0.3 to 7.3 ± 1.4 mg/dL, sCr decreasing from 0.9 ± 0.1 to 0.6 ± 0.1 mg/dL, and the SUA/sCr ratio rising from 7.0 ± 0.4 to 12.1 ± 7.6. The eGFR improved across quintiles in both sexes, increasing from 70.1 ± 6.8 to 111.9 ± 29.4 mL/min/1.73 m² in men, and from 67.4 ± 5.7 to 109.8 ± 155.0 mL/min/1.73 m² in women. Mean age was comparable across quintiles, around 59 years in men and 66 years in women, while the CCI varied only slightly between 0.3 and 0.5.

**Table 3.**
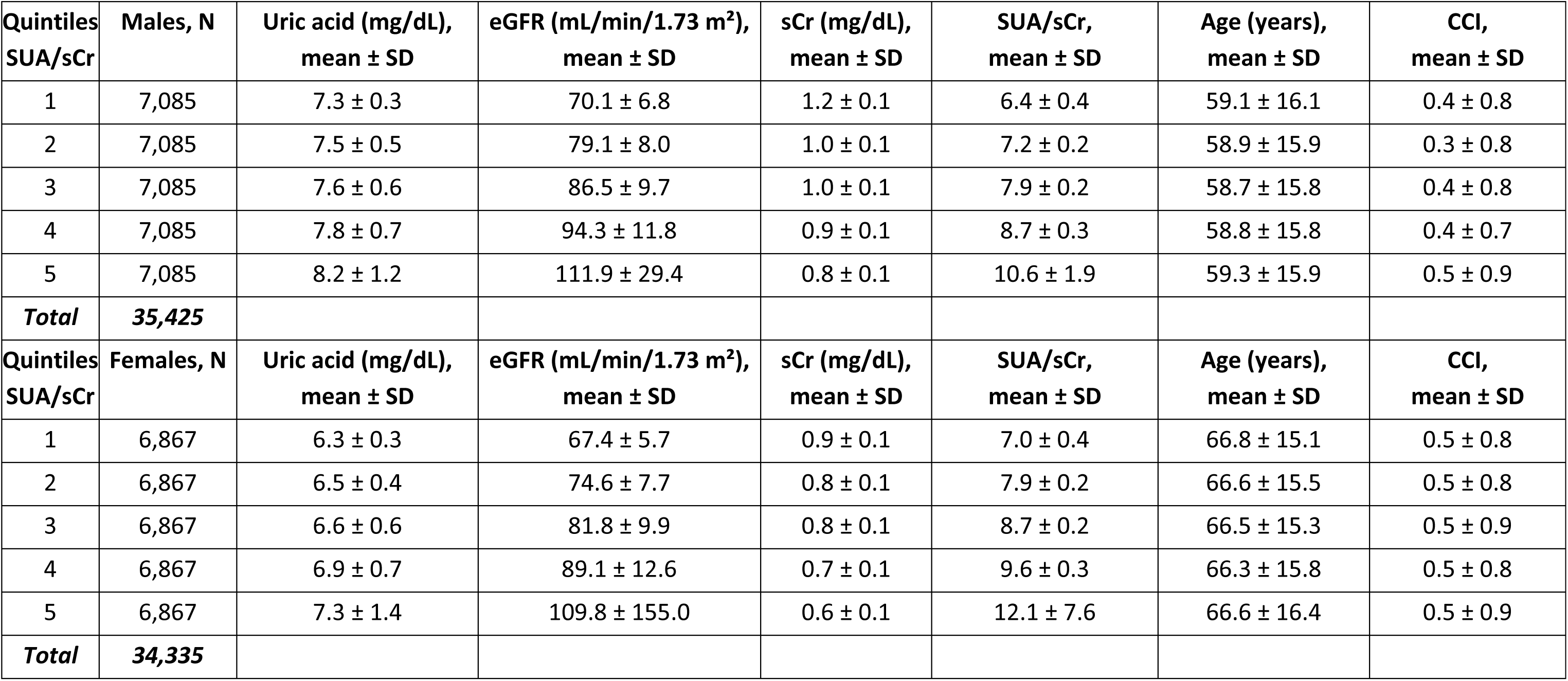
Demographic, biochemical, and clinical characteristics of patients with SUA levels above the upper reference value (7 mg/dL for males and 6 mg/dL for females).

| Quintiles<br>SUA/sCr | Males, N | Uric acid (mg/dL),<br>mean ± SD | eGFR (mL/min/1.73 m<sup>2</sup>),<br>mean ± SD | sCr (mg/dL),<br>mean ± SD | SUA/sCr,<br>mean ± SD | Age (years),<br>mean ± SD | CCI,<br>mean ± SD |
| --- | --- | --- | --- | --- | --- | --- | --- |
| 1 | 7,085 | 7.3 ± 0.3 | 70.1 ± 6.8 | 1.2 ± 0.1 | 6.4 ± 0.4 | 59.1 ± 16.1 | 0.4 ± 0.8 |
| 2 | 7,085 | 7.5 ± 0.5 | 79.1 ± 8.0 | 1.0 ± 0.1 | 7.2 ± 0.2 | 58.9 ± 15.9 | 0.3 ± 0.8 |
| 3 | 7,085 | 7.6 ± 0.6 | 86.5 ± 9.7 | 1.0 ± 0.1 | 7.9 ± 0.2 | 58.7 ± 15.8 | 0.4 ± 0.8 |
| 4 | 7,085 | 7.8 ± 0.7 | 94.3 ± 11.8 | 0.9 ± 0.1 | 8.7 ± 0.3 | 58.8 ± 15.8 | 0.4 ± 0.7 |
| 5 | 7,085 | 8.2 ± 1.2 | 111.9 ± 29.4 | 0.8 ± 0.1 | 10.6 ± 1.9 | 59.3 ± 15.9 | 0.5 ± 0.9 |
| **Total** | **35,425** |  |  |  |  |  |  |
| Quintiles<br>SUA/sCr | Females, N | Uric acid (mg/dL),<br>mean ± SD | eGFR (mL/min/1.73 m<sup>2</sup>),<br>mean ± SD | sCr (mg/dL),<br>mean ± SD | SUA/sCr,<br>mean ± SD | Age (years),<br>mean ± SD | CCI,<br>mean ± SD |
| 1 | 6,867 | 6.3 ± 0.3 | 67.4 ± 5.7 | 0.9 ± 0.1 | 7.0 ± 0.4 | 66.8 ± 15.1 | 0.5 ± 0.8 |
| 2 | 6,867 | 6.5 ± 0.4 | 74.6 ± 7.7 | 0.8 ± 0.1 | 7.9 ± 0.2 | 66.6 ± 15.5 | 0.5 ± 0.8 |
| 3 | 6,867 | 6.6 ± 0.6 | 81.8 ± 9.9 | 0.8 ± 0.1 | 8.7 ± 0.2 | 66.5 ± 15.3 | 0.5 ± 0.9 |
| 4 | 6,867 | 6.9 ± 0.7 | 89.1 ± 12.6 | 0.7 ± 0.1 | 9.6 ± 0.3 | 66.3 ± 15.8 | 0.5 ± 0.8 |
| 5 | 6,867 | 7.3 ± 1.4 | 109.8 ± 155.0 | 0.6 ± 0.1 | 12.1 ± 7.6 | 66.6 ± 16.4 | 0.5 ± 0.9 |
| **Total** | **34,335** |  |  |  |  |  |  |

Among patients with SUA levels above the upper reference threshold, two-year mortality progressively increased across SUA/sCr ratio quintiles in both sexes (Figure 3A–B). In males, mortality showed a slightly rising trend in the first three quintiles, and rose from 5.5% in the first quintile, then sharply to 11.3% in the fifth.(check) In females, the pattern was similar, with a modest rise from 6.3% to 7.0% within the third quintile, and then a more marked rise to 9.6% in the fourth and 15.4% in the fifth. Two-year CV event rates also increased in both sexes (Figure 3C–D). In men, the proportion of events increased from 14.3% in the lowest quintile to 20.0% in the highest. In women, CV events rose steadily from 11.5 in the first quintile to 18.2% in the fifth.

**Figure 3.**
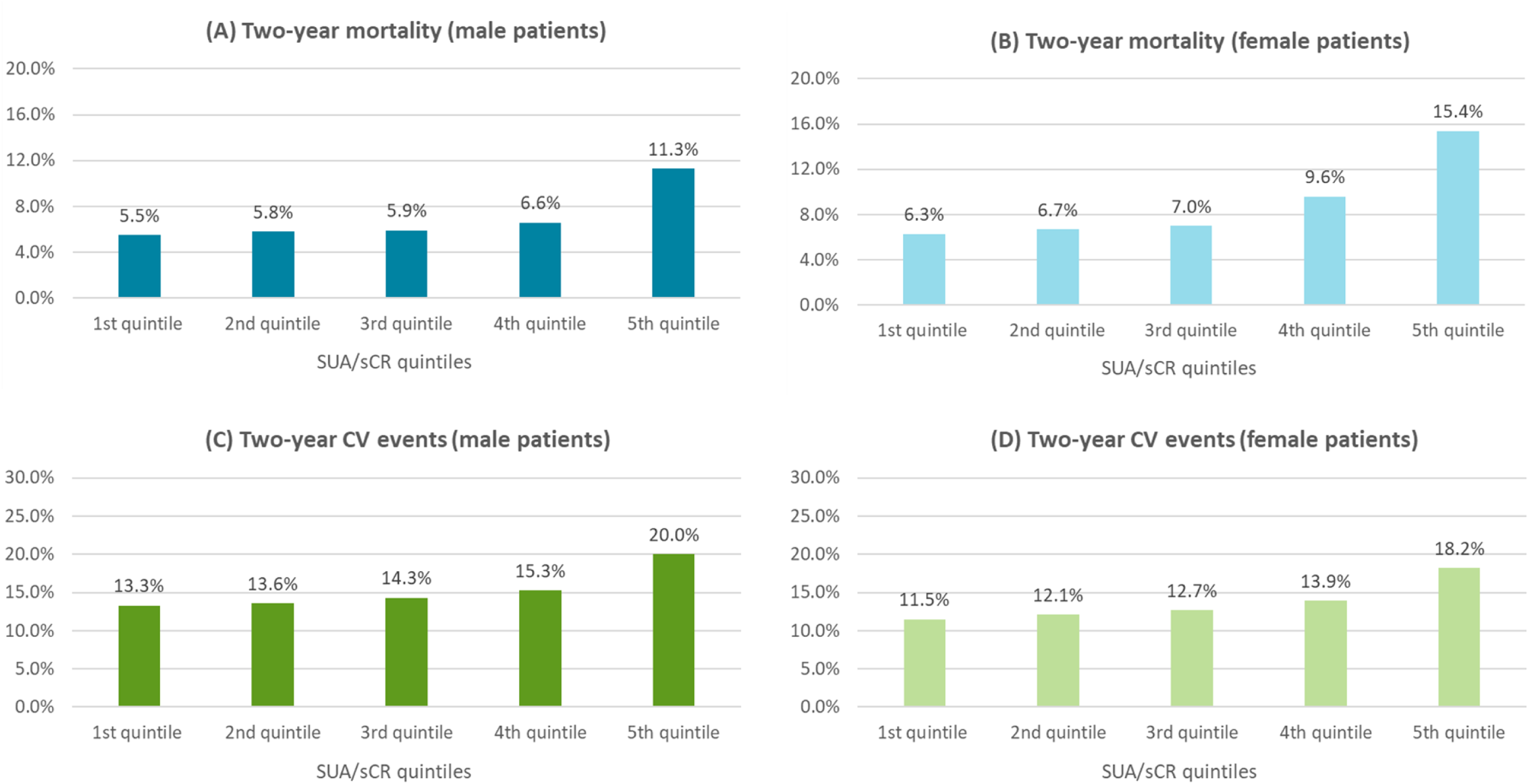
Two-year mortality in males **(A)** and in females **(B)**; CV events in males **(C)** and in females **(D)** divided by SUA/sCr quintiles in patients with SUA levels above the upper reference value (7 mg/dL for males and 6 mg/dL for females).

## Discussion

This large-scale, real-world analysis provides new insights into the prognostic role of the uric acid in an unselected Italian population using administrative and laboratory health databases. Our findings confirm and expand previous evidence on the relationship between SUA/sCr and the risk of all-cause mortality and CV events, particularly in individuals with hyperuricemia.

A key finding of this study is the progressive increase in both mortality and CV events within two years from the first SUA measurement beginning with the third SUA/sCr quintile and becoming more pronounced in the fourth and fifth. These results are consistent with and extend the hypothesis previously reported by Casiglia and colleagues,^17^ who identified a SUA/sCr threshold (>5.35 mg/dL) above which mortality risk increased in hypertensive individuals with preserved renal function. While the original URRAH data were based on smaller, highly characterized cohorts with long-term follow-up, our analysis involved nearly 150,000 individuals per quintile and reflects routine clinical practice across nine Italian regions, offering broader generalizability.

The rise in two-year mortality and CV events observed was particularly evident in patients with SUA above reference values. In this subgroup, event rates rose progressively across SUA/sCr quintiles in both sexes, with sharper increases in the upper quintiles. The results support the idea that the reason for the increased mortality and CV morbidity observed in the general population in patients falling into the first quintile is actually due to the unfavourable prognostic role of underlying clinical conditions leading to low levels of serum urate. These results reinforce the importance of integrating SUA with a renal function marker like sCr. Since sCr is inversely related to glomerular filtration, adjusting SUA for sCr offers a more nuanced picture of net urate load and excretory capacity. The SUA/sCr ratio thereby provides a dual signal: one representing the metabolic burden of uric acid and the other reflecting potential renal inefficiency.

Although our study is observational and based on administrative data, several strengths enhance its validity. First, the use of real-world data from a large, unselected population strengthens the external validity of the findings. Unlike research cohorts limited to specific subgroups (e.g., hypertensives or diabetics), our study included individuals aged 18 to 95 years, making the results applicable to a broad clinical setting. Second, the large sample size, roughly 740,000 individuals with normal renal function, offers unmatched statistical power and allows for robust stratification by SUA/sCr quintiles. Third, the predictive value of SUA/sCr was confirmed over a relatively short follow-up (2 years), contrasting with earlier studies that relied on 10–20 year horizons.^28-30^ This short-term predictive capacity enhances the clinical usefulness of SUA/sCr in identifying patients who may benefit from early preventive strategies.

In a nutshell, this study confirms the prognostic value of the SUA/sCr ratio for short-term mortality and CV risk in adults with preserved renal function. The association was strongest in individuals with elevated SUA, supporting the notion that the ratio reflects the combined impact of uric acid burden and renal excretory function. These findings align with mechanistic evidence linking hyperuricemia to endothelial dysfunction, oxidative stress, and inflammation.^31-35^ The clinical relevance of SUA/sCr has also been demonstrated in high-risk populations, including patients with diabetes^18^ and chronic kidney disease.^36,37^

Notably, when analyses were restricted to individuals with SUA levels above the sex-specific reference thresholds, the association remained monotonic and robust. This eliminates potential confounding from secondary hypouricemia and strengthens the argument for using SUA/sCr as a risk marker in clinical practice. The ability of this biomarker to stratify risk independent of detailed clinical data makes it especially useful for population-level surveillance and early intervention strategies.

Several strengths enhance the relevance of our findings. The SUA/sCr ratio proved meaningful predictive value over a short-term (2-year) horizon, distinguishing it from many prior studies requiring longer follow-up to detect CV outcome differences. Moreover, the ratio retained its prognostic performance in a large, unselected population despite the absence of granular clinical or lifestyle data. This highlights its potential utility in routine care and administrative health systems, where comprehensive data may not be available.

Nonetheless, some limitations must be acknowledged. First, our dataset lacked clinical and lifestyle variables such as smoking, alcohol consumption, blood pressure, heart rate, and anthropometric details. These unmeasured confounders, common in administrative databases, could influence outcomes. To mitigate this, we included the CCI to capture overall comorbidity, though it does not specifically reflect CV risk. Another limitation is the lack of detailed information on concomitant therapies across SUA/sCr quintiles. Medications such as diuretics, SGLT2 inhibitors, and urate-lowering agents can affect both SUA levels and clinical outcomes. Although individuals already receiving urate-lowering treatments were excluded, the absence of therapy-specific data may have limited our ability to detect treatment-related modifiers. Future studies incorporating prescription registries could provide deeper insights into these potential confounders.

Additionally, individuals in the lowest SUA/sCr quintile may have had low SUA due to secondary causes such as malnutrition, chronic inflammation, or malignancy. These conditions may have contributed to the higher mortality observed in this group. However, when the analysis excluded individuals with SUA below the reference range, the association between higher SUA/sCr and mortality remained consistent, suggesting that the observed effect is not solely driven by reverse causality or underlying illness.

In conclusion, the SUA/sCr ratio emerges as a valuable, independent predictor of short-term mortality and CV risk in adults with preserved renal function. Its predictive value, even in the absence of traditional confounders, highlights its potential utility in clinical and public health settings. Future research should focus on elucidating the biological basis of extreme SUA/sCr values, exploring therapy patterns across quintiles, and determining whether targeted interventions to modify the ratio can improve outcomes.

## Data Availability

The data supporting the findings of this article are available at aggregated level from the authors upon reasonable request and with permission of the participating healthcare entities. Requests to access should be directed to corresponding author.

## Nonstandard Abbreviations and Acronyms

ATC: Anatomical Therapeutic Chemical
CCI: Charlson Comorbidity Index
CKD: Chronic Kidney Disease
CV: Cardiovascular Disease
DRG: Diagnosis-Related Group
eGFR: Estimated Glomerular Filtration Rate
ICD-9-CM: International Classification of Diseases, Ninth Revision, Clinical Modification
INHS: Italian National Health System
SD: Standard Deviation
sCr: Serum Creatinine
SUA: Serum Uric Acid
SUA/sCr: Serum Uric Acid to Serum Creatinine ratio
URRAH: Uric Acid Right for Heart Health

